# Curation and description of a blood glucose management and nutritional support cohort using the eICU collaborative research database

**DOI:** 10.1101/2023.04.20.23288845

**Authors:** Oisin Fitzgerald, Oscar Perez-Concha, Blanca Gallego-Luxan, Lachlan Rudd, Louisa Jorm

## Abstract

Freely available electronic medical record (EMR) data collections have transformed data science and observational research in critical care medicine. Descriptive characterisation of these data collections can aid in highlighting variation in clinical practice and patient outcomes across Intensive Care Units (ICUs). Glycaemic control and nutritional management are important aspects of patient management in the ICU. Blood glucose on admission has a well-known U-shaped relationship with mortality and morbidity, with both hypo- and hyper-glycemia being associated with poor patient outcomes. The importance of nutritional support has been highlighted in critical care guidelines. However, both areas have open research questions and highly variable clinical practices that observational data may help highlight and inform. To aid in this research, we curated a database of patients using the eICU collaborative research data (eICU-CRD), which we describe in the current paper, focusing on patient blood glucose, insulin therapy and enteral nutrition. The eICU-CRD is derived from a telehealth EMR covering 208 United States hospitals from 2014-2015. In addition to descriptive statistics and graphical analysis, we highlight any limitations in data quality. Our results are in line with previous research suggesting the eICU-CRD cohort is of lower illness severity than the average ICU patient cohort and so receive less invasive interventions. Examinations of data missingness revealed issues with medication orders and non-reporting of nutrition by several hospitals. Overall, with care around missingness we believe the eICU-CRD to be a valuable resource in evidence generation for critical care research.

## Introduction

Freely available electronic medical record (EMR) data collections have transformed data science and observational research in critical care medicine Sauer et al. (2022). The MIMIC-III database (Johnson et al., 2016) is perhaps the most well-known and as of December 2022 has been cited more than 4,000 times and extensively used in research areas such as machine learning in medicine (Shillan, Sterne, Champneys, & Gibbison, 2019). Alternative data sources include the eICU collaborative research database (eICU-CRD) (Pollard et al., 2018), the Amsterdam University Medical Center database (AmsterdamUMCdb) (Thoral et al., 2021), the High time-resolution intensive care unit dataset (HiRID) (Faltys et al., 2021) and newer versions of MIMIC (Johnson et al., 2020). Given this increasing choice, researchers have sought to characterise the patient populations and data quality of the various datasets (O’Halloran, Kwong, Veldhoen, & Maslove, 2020; Sauer et al., 2022), and create curated datasets geared towards particular clinical applications such as glycaemic control (Arévalo et al., 2021). In this article we provide a descriptive introduction to the use of eICU-CRD, a database constructed from the Philips eICU program, a critical care telehealth service, for research relating to blood glucose management and nutritional support in the ICU.

Glycaemic control is a core aspect of patient management in the ICU. Blood glucose on admission has a well-known U-shaped relationship with mortality and morbidity, with both hypo- and hyper-glycaemia being associated with poor patient outcomes (Siegelaar et al., 2010). As a result, up to 40-90% of ICU patients receive insulin, with the upper bound applicable to patients with longer ICU stays and tighter blood glucose targets (Fitzgerald et al., 2021; Nice-Sugar Study Investigators, 2009; van Steen, Rijkenberg, van der Voort, & DeVries, 2019). Generally clinical guidelines for glycaemic control (e.g. Qaseem, Chou, Humphrey, Shekelle, and Physicians (2014)) are based on a series of trials that culminated in the NICE-SUGAR study (Nice-Sugar Study Investigators, 2009), a multicentre study demonstrating that tight glycaemic control (a target of 80-140 mg/dL) did not improve patient outcomes compared to moderate control (<180 mg/dL). However, there remain open questions around the impact of glucose variability (Ceriello & Ihnat, 2010), the potential for more personalised glycaemic targets (Sechterberger et al., 2013), and the potential for an “artificial pancreas” via control algorithms (Chase et al., 2006), which openly available EMR collections such as eICU-CRD can play a role in answering.

Nutritional support in the ICU is increasingly recognised as an important issue, with variation in degree and timing of feeding associated with patient mortality and morbidity (Marik, 2014; Preiser et al., 2021). It is also clearly related to glycaemic control, with enteral and parental nutrition (PN) parameters predictive of future blood glucose values (Fitzgerald et al., 2021). Nevertheless, many blood glucose protocols are solely based on previous blood glucose and insulin measures (Krikorian, Ismail-Beigi, & Moghissi, 2010; Wilson, Weinreb, & Hoo, 2007). As with glycaemic control, there exist open questions around optimal nutritional support, such as optimal total calories and protein and timing of nutrition (Preiser et al., 2021). For example, clinical guidelines for nutrition in the ICU increasingly recommend early (24-48 hours) initiation of enteral nutrition (EN) to reduce morbidity and mortality (McClave et al., 2016; Singer et al., 2019). However, there remain some concerns around the standard of evidence supporting these recommendations (Padilla et al., 2019). While studies of nutritional support using observational data such as eICU-CRD may not necessarily be considered a high-quality source of evidence, the large size and immediate availability will help in goals such as selecting target population who may benefit the most from clinical trials (such as in Zheng et al. (2022)).

Clearly, glycaemic control and nutritional support are areas of critical care in which further research is warranted. High quality data are of particular importance for research investigating longitudinal relationships between patient characteristics, patient management and patient outcomes with the aim of personalising care (e.g. development of reinforcement learning algorithms). Details of the timing and nature of an action or decision (for example, insulin rate/dose) are required in order to model its effects. O’Halloran et al. (2020) previously published a high-level description of the patient population and data quality of the eICU-CRD. Their main findings were: 1) the patient population has a lower illness severity (based on mortality rate, length of stay and interventions) than the average ICU in the USA; 2) vital signs are generally well recorded but certain lab results commonly used in severity scores have low reporting rate; and 3) complex interventions (e.g., dialysis, intubation) have unclear prevalence due to heterogeneity in how they are recorded in the source database. We aim to extend this analysis to glycaemic control and nutritional support, characterising aspects of the data such as patient blood glucose, insulin therapy and enteral and parental nutrition using descriptive statistics and graphical analysis and highlighting any limitations in data quality.

## Methods

### Data source

Data for this study were sourced from the eICU collaborative research database (eICU-CRD) open access critical care database, de-identified to conform with the Health Insurance Portability and Accountability Act (HIPAA). eICU-CRD is a large multi-center critical care database holding data associated with 200,859 ICU stays admitted at 208 hospitals across the United States between 2014 and 2015 (Pollard et al., 2018).

### Data extraction, pre-processing, and analysis

We accessed the eICU-CRD database using Google BigQuery, performing the data curation steps using the open source dbt package (https://docs.getdbt.com/) a SQL based data modelling tool. The contents and schema of the resulting database are outlined in Appendix A. Following this step we imported the data into R version 4.2 (Team, 2013) using the bigrquery package (H Wickham & Bryan, 2018), with the data.table package (Dowle & Srinivasan, 2019) used for further data cleaning and summarisation. Data engineering and pre-processing steps are outlined below and in further detail in the associated code repository www.github.com/oizin/eicu-glucose-cohort.

The analysis was descriptive in nature, using counts, percentages, transformations, and summary statistics as appropriate. Given the skewed nature of many variables, continuous variables are reported as median and interquartile range unless otherwise stated. Graphical analysis was performed using ggplot2 (Hadley Wickham, 2011). All analyses were by ICU stay (rather than patient) unless stated otherwise. In cases where the “admission” value of a variable is reported this is first value recorded during a patient’s ICU stay. In contrast to some of the analyses reported in (O’Halloran et al., 2020) we excluded or set to missing (i.e. NA) clearly implausible values (e.g. patient weights above 500 kg), and attempted to transform variables (e.g. insulin infusions) to consistent scales where the data was recorded heterogeneously (which was the norm).

### ICU stay characteristics and outcomes

The *patient* and *icustay* tables were used to extract details of the ICU stay (admission and discharge date, ICU and hospital outcomes, and APACHE admission diagnosis, APACHE-IVa score (Zimmerman, Kramer, McNair, & Malila, 2006)), patient characteristics/demographic information (age, weight, diabetic status) and the ICU/hospital (medical/surgical/cardiac, teaching status and region).

### Blood glucose measurements

Blood glucose measurements were extracted from the *nurseCharting* and *lab* tables. Point of care (bedside) measurements were present in both tables, with *lab* additionally containing results that occurred as part of linkable lab results. Details of the measurement device were not recorded in the database but are presumably chemistry analysers for lab measurements and fingerstick glucometers (or in rare cases arterial blood gas analysers) for bedside measures, in line with common practice.

### Insulin

The data items in the *intakeOutput, infusionDrug, medication and treatment* tables were searched for any items related to insulin. The *medication* table contains active medication orders, with information on the product ordered (e.g., 100 units/mL of Aspart) or a product code, start and stop times, the route, dose, frequency, whether the insulin was to be given as required (PRN from the Latin “pro re nata” or “as required”) or at a set frequency and whether the insulin was an intravenous (IV) admixture. This data was recorded in a heterogenous manner, for example,”sc” or “s.c.” used to indicate subcutaneous (SC) insulin. During data cleaning, these data was converted into variables that indicated whether the insulin order was SC or IV, PRN, by the type of insulin (aspart, lispro, regular, glargine, or detemir - inferred from the product name), insulin analog rate of action (e.g., short or long), and by the implied daily frequency of treatment (for example, “t.i.d.” to 3, that is 3 times a day, from the Latin “ter in die”). Additionally, the character column dosage was converted to numeric column “dose”, or in some cases to “dose low” and “dose high” where (for example) “1-10 UNITS” was the value in the data.

The rate of IV insulin (units/hour), and infusion start times were extracted from the *infusionDrug* table. The *infusionDrug* table contains the IV insulin infusion start time, and either the rate or total insulin units given. This table does not contain an infusion end time column and so changes in the infusion (such as termination) need to be inferred by linking together all rows for a particular ICU stay and using the next event time to infer a change. Additional, similar information was extracted from the *intakeOutput* table, with the information from these two tables then combined.

### Intravenous dextrose

All distinct data items in the *intakeOutput, infusionDrug* and *medication* tables were assessed as to whether they indicated IV glucose (dextrose) intake. The infusion start/stop times and glucose content (0.45%, 5%, 10% or 50%) were extracted.

Importantly while *infusionDrug* and *intakeOutput* contain interventions performed, the *medication* table is orders, which are not necessarily administered, a note applicable to insulin and IV dextrose.

### Nutrition

All distinct data items in the *intakeOutput, infusionDrug, medication, treatment and nurseCare* tables were assessed as to whether they were related to non-specific oral nutritional intake (liquid or solid), oral intake of glucose, EN, PN, or other unspecified nutrition and categorised as such. The EN data items were further subcategorised into whether they indicated tube feeding of a specific product (e.g. Nutren 1.5), tube feeding of an unspecified product, intake of non-food (water, medication intake or flushing) or, other enteral related procedure. The start/stop time, and rates of EN and PN were extracted.

## Results

An overview of the ICU stays and patient characteristic found in the eICU-CRD database can be found in Table 1. We include additional results, including a breakdown of admission diagnosis and characterisation of hospital variability in the use of insulin and nutritional support in Appendix B. The data included 200,859 ICU stays across 166,355 patients, a mean of 1.2 ICU stays per individual. The median length of ICU stay was 1.6 days (IQR: 0.8; 3.0) with a hospital mortality rate of 9.0%. The majority (92%) of stays were over 6 hours with patient mortality accounting for 14% of those with stays under 6 hours. ICU stays by older and female patients resulted in higher hospital mortality rates (the mortality rates for those over and under 60 was 10.9% and 6.5%, and for females and males was 9.2% and 8.9%, respectively), with age at ICU admission being on average 2 years younger for males than females (64 and 66 years). Among the 90.1% of ICU stays with weight and height information available, 36.9% were for obese patients (BMI > 30 kg/m^2^), with the median patient BMI in the overweight range (27.5 kg/m^2^, IQR of 23.5-32.9 kg/m^2^). A higher BMI was a protective factor for hospital mortality (the so-called “obesity paradox” (Dickerson, 2013))(Table 1). One-fifth (19.6%) of ICU stays were for patients with diabetes, with a diagnosis of diabetes also associated with reduced hospital mortality (8.4% vs 9.8% for non-diabetics). Caucasian patients accounted for the majority of ICU stays (77.3%), while ICU stays by Hispanic patients had the highest mortality rate (9.9%).

**TABLE 1.** Overview of the eICU-CRD cohort. Note that due to missing outcome columns may not add to all.

| Continuous variables are median (IQR)<br>Categorical variables are count (%) | All | Hospital outcome<br>Survived | Died in hospital |
| --- | --- | --- | --- |
| *Stay characteristics* |  |  |  |
| ICU stays | 200,859 | 181,104 (90.2%) | 18,004 (9.0%) |
| ICU stays over 6 hours | 185,473 | 168,015 (90.6%) | 15,846 (8.5%) |
| Patients | 166,355 | 150,252 (90.3%) | 14,623 (8.8%) |
| Age (years) | 65 (53-76) | 64 (52-75) | 72 (61-81) |
| Gender |  |  |  |
| Male | 108,379 | 97,875 | 9,608 (8.9%) |
| Female | 92,303 | 83,097 | 8,372 (9.1%) |
| Missing |  |  |  |
| BMI (kg/m<sup>2</sup>) | 27.5 (23.5-32.9) | 27.6 (23.6-33.0) | 26.6 (22.5-32.2) |
| Underweight (BMI < 18.5) | 8,204 | 6,984 (85.1%) | 1,135 (13.8%) |
| Normal (BMI 18.5-25) | 55,479 | 49,539 (89.3%) | 5,450 (9.8%) |
| Overweight (BMI 25-30) | 54,643 | 49,761 (92.0%) | 4,409 (8.0%) |
| Obese (BMI > 30) | 69,324 | 63,340 (91.4%) | 5,433 (7.8%) |
| *Diabetic* |  |  |  |
| Yes | 39,308 | 35,698 (90.8%) | 3,282 (8.3%) |
| No | 131,869 | 117,834 (89.4%) | 12,811 (9.7%) |
| Missing | 29,682 | 27,572 (92.9%) | 1,911 (6.4%) |
| *Ethnicity* |  |  |  |
| Caucasian | 155,285 | 140,030 (90.2%) | 13,934 (9.0%) |
| African American | 21,308 | 19,268 (90.4%) | 1,865 (8.8%) |
| Hispanic | 7,464 | 6,692 (89.7%) | 736 (9.9%) |
| Asian | 3,270 | 2,911 (89.0%) | 310 (9.5%) |
| Native American | 1,700 | 1,550 (91.2%) | 138 (8.1%) |
| Other/unknown | 11,832 | 10,653 (90.0%) | 1,021 (8.6%) |
| *ICU type* |  |  |  |
| Mixed | 113,222 | 102,223 (90.3%) | 9,906 (8.7%) |
| Medical | 17,465 | 15,097 (86.4%) | 2,231 (12.8%) |
| Surgical | 12,181 | 10,996 (90.3%) | 1,124 (9.2%) |
| Cardiac (CCU/CTICU) | 43,540 | 40,025 (91.9%) | 3,515 (8.1%) |
| Neurological | 14,451 | 13,048 (90.3%) | 1,228 (8.5%) |
| *Admission type* |  |  |  |
| Non-operative | 143,620 | 128,453 (89.4%) | 15,167 (10.6%) |
| Operative (non-elective) | 3,666 | 3,296 (89.9%) | 370 (10.1%) |
| Operative (elective) | 30,577 | 29,591 (96.8%) | 986 (3.2%) |
| APACHE-IVa score | 51 (37; 68) | 49 (36; 64) | 84 (64; 108) |
| *Blood glucose measurements* |  |  |  |
| ICU stays with at least one measurement | 175,952 | 158,981 (90.4%) | 15,440 (8.8%) |
| Measures per day (median IQR) | 4.3 (2.7; 7.2) | 4.2 (2.7; 7.1) | 4.6 (2.9; 7.6) |
| Admission blood glucose (mg/dL) | 132 (109; 167) | 131 (108; 165) | 143 (113; 186) |
| Mean blood glucose (mg/dL) | 128 (108; 157) | 127 (107; 156) | 141 (117; 173) |
| Max blood glucose (mg/dL) | 161 (123; 224) | 158 (121; 219) | 198 (149; 273) |
| Blood glucose SD (mg/dL) | 24 (14; 40) | 24 (14; 39) | 32 (20; 50) |
| Any hyperglycaemic (>180 mg/dL) measure on day 1 | 59,703 | 51,989 (87.1%) | 7,214 (12.1%) |
| Any hypoglycaemic (<70 mg/dL) measure on day 1 | 9,936 | 8,027 (80.7%) | 1,819 (18.3%) |
| *Insulin therapy* |  |  |  |
| Any insulin | 74,464 | 66,181 (88.9%) | 7,693 (10.3%) |
| *Insulin route* |  |  |  |
| Intravenous (IV) | 23,506 | 20,689 (88.0%) | 2,693 (11.5%) |
| Subcutaneous (SC) | 58,023 | 51,947 (89.5%) | 5,690 (9.8%) |
| *Nutrition / IV dextrose* |  |  |  |
| *Route of intake* |  |  |  |
| Enteral (tube feeding) | 14,509 | 11,600 (79.9%) | 2,741 (18.9%) |
| Parenteral | 5,676 | 4,627 (81.5%) | 993 (17.5%) |
| IV dextrose | 64,839 | 57,333 (88.4%) | 7,161 (11.0%) |
| *Patient outcomes* |  |  |  |
| Died in ICU | 10,941 | 0 (0.0%) | 7,315 (66.9%) |
| Length of ICU stay (days) | 1.6 (0.8; 3.0) | 1.5 (0.8; 2.9) | 2.0 (0.7; 4.8) |
| Length of hospital stay (days) | 5.5 (2.9; 10.0) | 5.5 (3.0; 9.9) | 5.3 (1.9; 11.9) |

\*1,751 ICU stays are missing their hospital outcome

Most ICU admissions were to a mixed medical-surgical ICU (56.4%) with specialist cardiac or neurological ICUs accounting for 28.9%. Among the operative ICU stays, 10.7% were elective, with the elective surgeries experiencing a far lower hospital mortality rate (3.2%) than non-elective surgical stays (10.1%). The most common APACHE admission diagnosis was some form of sepsis (11.5%) with these ICU stays associated with an above average hospital mortality rate of 16.5%. A similar proportion of patients (11.4%) had no reported admission diagnosis.

Based on the APACHE admission diagnosis, just under 3% of ICU admissions were primarily for derangements in blood glucose homeostasis. These admission diagnoses included diabetic ketoacidosis (DKA) and hyperosmolar hyperglycaemic coma (HHC) which were present in 2.4% and 0.2%. In line with expectations, the HHC group had higher blood glucose levels at ICU admission, with a median of 642 mg/dL (IQR: 468-881 mg/dL) for HHC stays and 456 mg/dL (IQR: 331-599 mg/dL) for DKA. The ICU length of stay and hospital outcomes for DKA and HHC were comparable, with respective median ICU length of stay of 25 hours (IQR: 18-43 hours) and 29 hours (IQR: 19-39 hours), and in-hospital mortality of 1.2% and 0.7%. A further 0.25% of patients were admitted primarily for hypoglycaemia. These admissions had a median blood glucose of 77 mg/dL (IQR: 45-123 mg/dL), a median 32 hour (IQR: 19-56 hour) stay in ICU and a 5.0% hospital mortality rate.

Insulin therapy was prescribed or administered in 31.0% of ICU stays. Patient blood glucose at admission for these ICU stays was typically h23 mg/dL higher than for patients who were not treated with insulin and patients treated with insulin during their stay were more likely to be diabetic than the non-treated population (55.1% vs. 22.9%). The most common route of insulin administration was SC injection (88.8% of cases) rather than IV infusion.

### Blood glucose measurements

Using the *bedside* and *lab* tables we extracted 2,351,977 bedside and 823,513 lab measurements after ensuring the measurement occurred within an ICU stay (or at most 12 hours prior to ICU admission) and accounting for duplication of measurements between and within tables. There were 2 hospitals with no blood glucose measurements, although these only accounted for 36 ICU stays. Among the hospitals with reported blood glucose measurements, 95.3% of admissions with an ICU stay of at least 12 hours had at least one blood glucose measurement, with a median number of bedside measurements per day of 4.3 (IQR 2.7-7.2) (Table 1). Factors that were related to more frequent blood glucose measurement were insulin prescription (5.5 measures per day (IQR: 3.8-10.5)), diagnosis of diabetes (5.2 measures per day (IQR: 3.9-9.3 hours)), a hypoglycaemic measurement on day 1 of the ICU stay (6.3 measures per day (IQR: 4.1-11.0)) and a hyperglycaemic measurement on day 1 of the ICU stay (5.3 measures per day (IQR: 3.8-10.1 hours)). From Figure 1D we see that the distribution of time between bedside blood glucose measurements is bimodal, with a tendency for blood glucose measurements to occur approximately either every 1 hours or every 4 hours, with a long tail of infrequent measurements. This is presumably a combination of the influence of standard practises (e.g., rounds and standard insulin injection times) on the time of measurement (the most common hours of measurement were 16:00-17:00, 22:00-23:00 and 04:00-05:00), and above outlined patient specific factors influencing the frequency blood glucose measurement.

**Figure 1.**
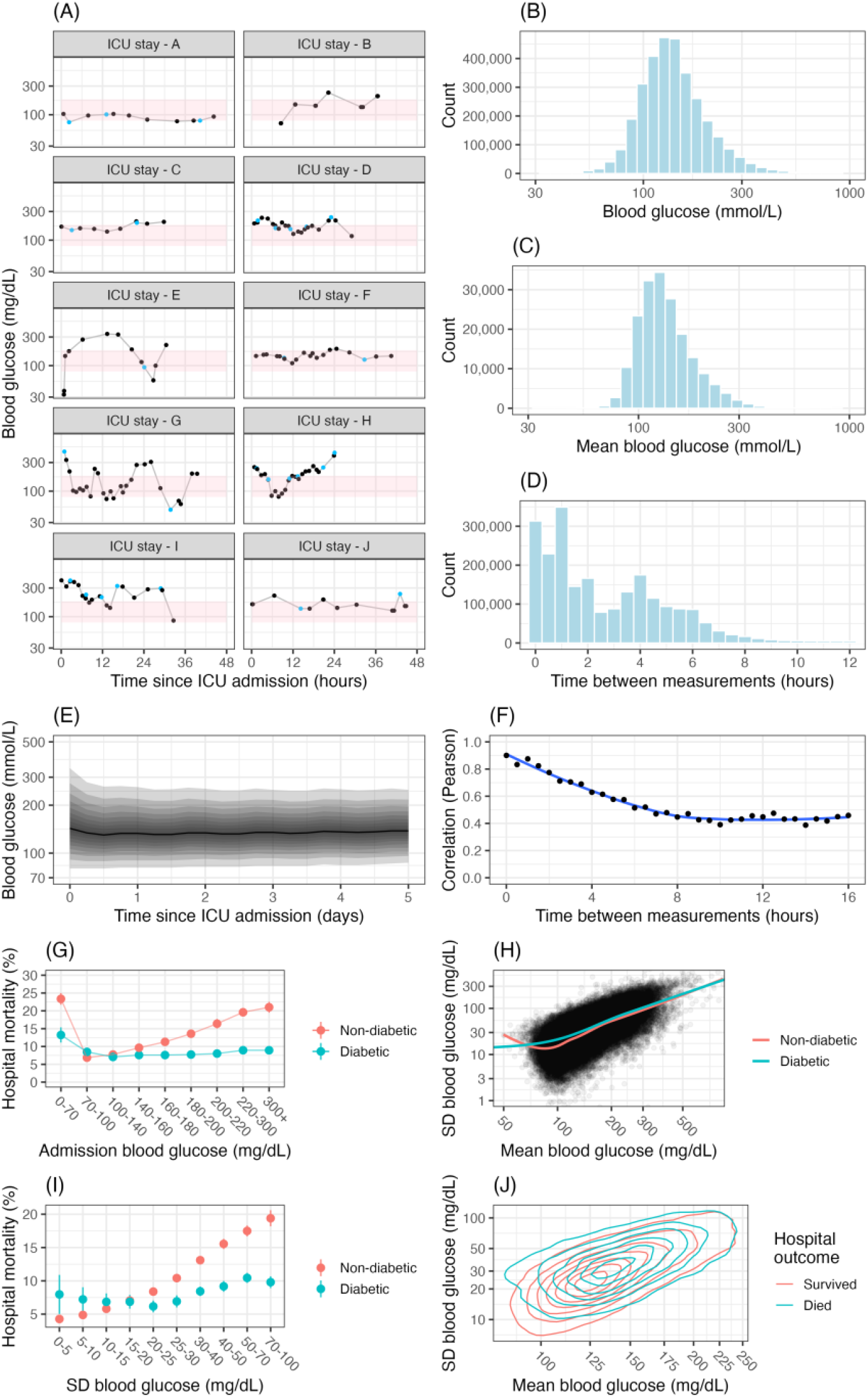
Characterising blood glucose in eICU-CRD. A) Sample blood glucose trajectories of 10 different patients during their ICU stay. The pink shaded region is 80-180 mg/dL. Black points are bedside blood glucose measurements while blue coloured points are laboratory measurements. B) The distribution of bedside blood glucose measurements. C) The distribution of laboratory blood glucose measurements. D) The distribution of the time between blood glucose measurements. E) The distribution of bedside blood glucose measurements over time. The solid line is the median. The shaded regions are the percentiles between the 2.5^th^ and 97.5^th^ percentiles incrementing by 5. F) The correlation between blood glucose measurements over time. G) Hospital mortality by admission blood glucose stratified by diabetic status. H) Scatter plot of mean and standard deviation (SD) of blood glucose across ICU stay with added smoothing line I) Hospital mortality by blood glucose SD across ICU stay J) 2D density plot of mean and standard deviation (SD) of blood glucose across ICU stay stratified by hospital outcome

As seen in Table 1, the proportion of patients experiencing hypoglycaemic and hyperglycaemic episodes on day 1 of an ICU stay was 29.7% and 4.9%, respectively. As expected using average blood glucose on day 1 of the ICU stay these figures reduced to 17.1% and X%. The severity of both hypoglycaemia and hyperglycaemia tended to decrease following the initial 24 hours of ICU stay and remain stable thereafter (Figure 1E). The distribution of all blood glucose measurements and the average blood glucose during ICU stays are shown in Figure 1B and 1C. As expected, the average blood glucose distribution is narrower, with a reduction in observations in the hypoglycaemic and hyperglycaemic ranges.

In patients who died in hospital, blood glucose tended to be both higher and show greater variability (Figures 1G-J). On average, these patients had a 14 mg/dL higher average blood glucose and 8 mg/dL higher average standard deviation. In non-diabetics, blood glucose had a U-shaped risk curve, with both hypoglycaemia and hyperglycaemia associated with increased mortality (Figure 1G). In diabetics the hyperglycaemic portion of this risk profile was greatly attenuated, with the hypoglycaemic portion of the risk curve generally shifted rightward compared to non-diabetics (Figure 1G). The inter-patient blood glucose variability is visible in Figure 1A where we see patients with relatively well controlled blood glucose (e.g., ICU stay - A) and patients with high average (e.g., ICU stay - D) and high variability (e.g., ICU stay - G) blood glucose. Within patients, blood glucose measurements trended towards a moderate degree of regularity over time, with the correlation between measurements plateauing at 0.5 after 10 hours between measurements (Figure 1F).

### Insulin management

Insulin orders and infusions can be found in the *medication, infusionDrug, intakeOutput* and *treatment* tables. There were 7 hospitals (3.4%), accounting for 308 ICU stays, where no patient was indicated as having received insulin. There was considerable heterogeneity in the completeness of the data between these tables. The *medication* table had a low rate of data completeness with 20.7% of rows having a perfectly complete record (defined as the availability of type, dose and frequency). The *infusionDrug* table was largely complete with 99% of insulin events containing a complete set of information (defined as the availability of insulin rate and start time). The *treatment* table contained information on whether the patient initiated an insulin infusion or received a short or long acting insulin injection at a particular time, with no information on the dose or rate.

A patient received IV insulin during 23,506 ICU stays (Table 1). Unfortunately, 81.0% of IV insulin orders in the *medication* table did not have any dose information and generally just noted the product ordered (e.g., “insulin regular human 100 unit/ml”) which did not allow for deterministic calculation of an infusion rate. Of the 4,006 ICU stays where insulin infusion details were recorded in *intakeOutput* half (48.3%) were additionally reported in the *infusionDrug* table. As such we largely concentrate on the 12,918 ICU stays in the *infusionDrug* table in characterising insulin infusion therapy in the eICU-CRD due to more consistent data (*intakeOutput* contained a mix of units). For these ICU stays the median infusion rate was 2.4 units/hour (IQR: 1.0-5.0), albeit with considerable variation (Figure 3B). Here we have assumed for the 26% of cases where the units (part of the drugname column) are mL/hr the solution contains 1 unit per mL. Diabetics were more likely to receive a higher dose, with a median rate of 3.0 units/hour (IQR: 1.8-6.0).

**Figure 3.**
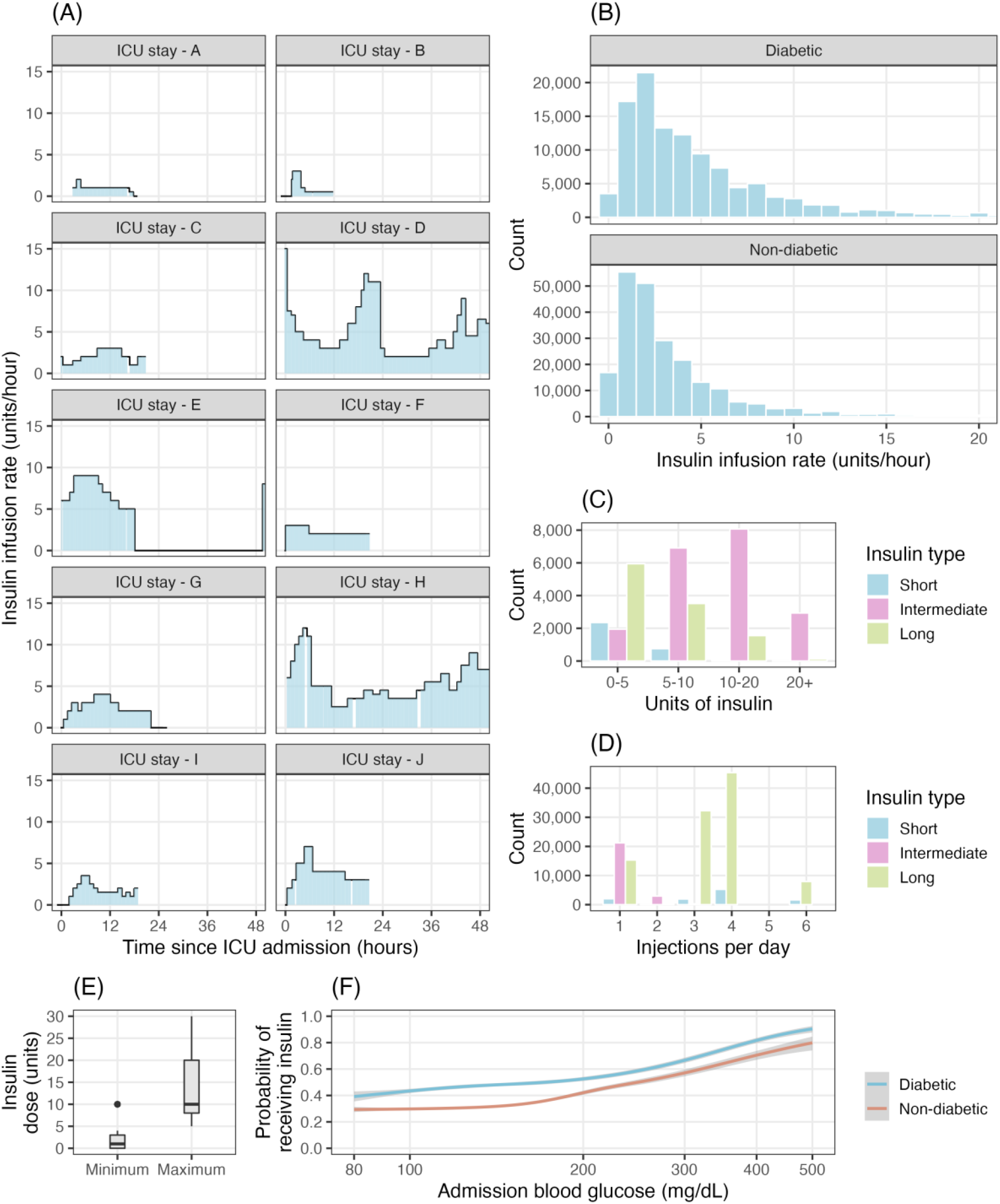
Characterisation of insulin therapy in the eICU-CRD. A) Sample insulin infusion rates of 8 ICU stays. B) Distribution of insulin infusion rate stratified by diabetic status. C) SC insulin dosage by type of insulin. D) SC insulin injections per day by type of insulin E) SC insulin minimum and SC dosage. F) Probability of receiving insulin on day 1 of an ICU stay by blood glucose level on admission and diabetic status

For the 58,023 ICU stays where SC insulin was ordered, a total of 192,033 active orders were available (from the *medication* table), with 15.5% containing full information on dose, insulin type and frequency. The most common order was for a short acting insulin (65.8%) followed by long acting insulin (11.6%), with short acting insulin typically a low dose (0-5 units) and frequency of 3-4 times a day while long acting orders were most typically at least 10 units and once per day (Figure 3C and 3D).

As expected, higher blood glucose and diagnosed diabetes were associated with an increased likelihood of some form of insulin therapy (Figure 3F). The form of insulin therapy used varied by hospital with small hospitals (<100 beds) less likely to use infusion (8% of patients prescribed insulin), compared to larger hospitals (20% of patients prescribed insulin).

### Intravenous dextrose

Similar to insulin, details of IV dextrose orders and administrations can be found in the *medication, infusionDrug* and *intakeOutput* tables. As above the *medication* table contains orders which are not guaranteed to be administered, however for simplicity we group all information across the tables for the purpose of this analysis and refer to “IV dextrose orders”. Across these tables it was possible to determine the concentration of the IV dextrose for 100% of orders using the product name (or equivalent variable). IV dextrose was ordered in 64,839 stays. The most common order was for 5% dextrose (59.7%). A large percentage (24.1%) of ICU stays had a 50% dextrose order (35.4% of orders), with these patients generally having a lower minimum blood glucose level compared to patients who did not have a 50% dextrose order (92 mg/dL vs 99 mg/dL). Amongst the ICU stays indicated as receiving IV insulin, 78.3% were indicated as having an IV dextrose order during their stay, most commonly 5% dextrose (53.1%), followed by 50% dextrose (42.3%). There were 77 hospitals, accounting for 34,813 ICU stays, where no ICU stay was indicated as having received IV dextrose.

### Oral, enteral and parenteral nutrition

Information on nutritional support, including oral (eating or drinking), EN or PN feeding, was found largely in the *intakeOutput* table (1,377,059 events across 72,346 ICU stays). To a far lower degree some indications of PN were found in the *infusionDrug* and *medications* table (13,161 events across 656 ICU stays). The *treatment* table contains an indication of PN and EN for 1,026 and 3,744 ICU stays, with the *nurseCare* table containing an indication of PN and EN for 214 and 2,163 ICU stays. For the latter two tables only the event name (e.g., “enteral” or “regular diet”) and the time is available with no rates or quantities recorded. The presence of nutritional information varied widely across patients and hospitals. As seen in Figure 4A, patients were more likely to have reported nutritional support with an increasing length of ICU stay, with variation in the intake route by illness severity (Figure 4B). While the lack of patient data may be partially explained by the high rate of no feeding common in ICU (discussed below), there were 34 hospitals (16%), accounting for 10,318 ICU stays, containing no nutritional information at all. Additionally for 68,039 ICU stays across 99 hospitals, it could only be inferred the patient has some form of intake, with no quantity/volume recorded. Further, the remaining reported wide variation in the rates of nutritional support (Figure 4B).

**Figure 4.**
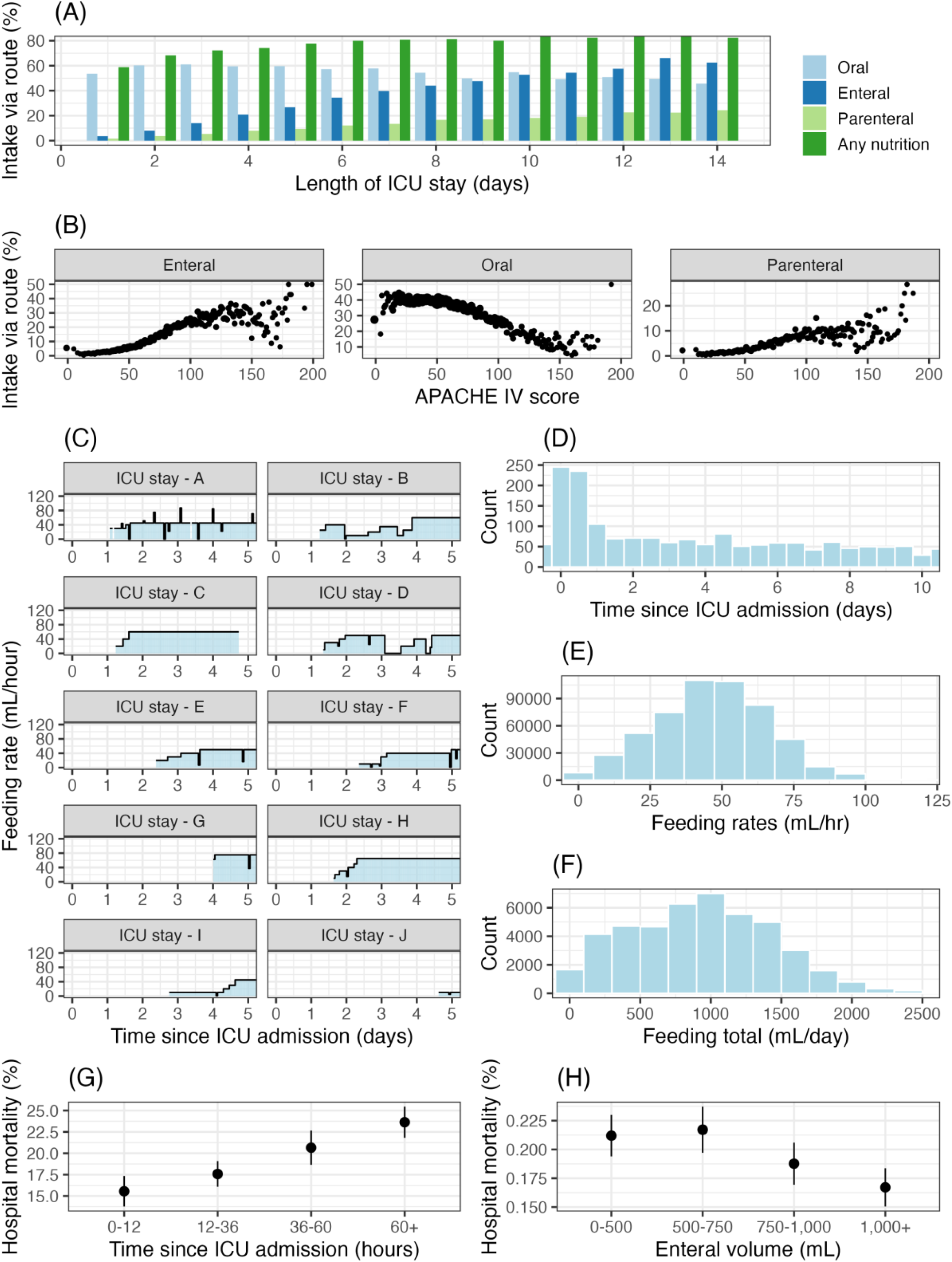
Charactering EN in the eICU-CRD. A) The percentage of ICU stays recorded as receiving EN, PN or oral nutritional intake by their length of ICU stay. Note this figure excludes hospitals where no nutritional information quantity is recordedl. B) Percentage of ICU stays receiving nutritional support via intake route by APACHE-IV score C) Sample EN feeding rate profiles. D) The distribution of EN starting times E) The distribution of EN feeding rates. F) The distribution of the EN total daily feeding. G) Hospital mortality by EN starting time H) Hospital mortality by average daily EN feeding volume

Amongst ICU stays receiving enteral nutritional support (for examples see Figure 4C) the median start time was 32 hours (IQR: 14-63 hours) after admission to ICU (see Figure 4D), with earlier initiation of EN associated with better hospital outcomes (Figure 4G). In a large percentage of cases the product used was not recorded (>26%), with Abbot Pivot and Nepro the most common recorded products (>9% of EN under different naming conventions). The median feeding rate was 45.0 mL/hr (IQR 30.0-60.0) (see Figure 4D). As seen in Figure 4E, assuming a 1.5-2.0 calorie per mL product was used, the majority (78.0-54.7%) of EN fed patients were likely to have caloric intakes under 2,000 calories, with 55.7-37.0% under 1,500 calories. As with earlier initiation, higher intake of EN was associated with better hospital outcomes (Figure 4H).

## Discussion

The goal of this paper was to support research into glycaemic control and nutritional support in the ICU through curating an eICU-CRD database and characterising the resulting cohort. The eICU-CRD is a de-identified release of data from the Philips telehealth program, and to the best of our knowledge is the only open access critical care telehealth EMR database. It is unique because it contains detailed information on ICU stays across multiple hospitals, with a large cohort of over 200,000 ICU stays. We have given a detailed analysis, including summary statistics, graphs and missingness analysis of the blood glucose, insulin therapy, and nutritional (IV dextrose, EN and PN) data that is often duplicated across several tables in the database, depending on how the originating hospital used the telehealth EMR. Overall, the eICU-CRD appears to be an excellent resource for research relating to glycaemic control and nutritional support. For certain research questions, in particular those requiring information on the timing and dosage of SC insulin, issues with missing information may limit the usefulness of the database, or require analysis of a smaller subset of hospitals with higher rates of data completion.

### Relationship to previous research

The eICU-CRD patient cohort has previously been noted to have a lower illness severity than the typical US ICU (O’Halloran et al., 2020), with a hospital mortality rate of 9.0%. In comparison, the commonly used MIMIC-III database, constructed from the ICUs in the Beth Israel Deaconess Medical Center (Boston), has a hospital mortality rate of 11.5%, more in line with the typical US ICU.

The use of insulin is comparable with other observational ICU EMR databases such as MIMIC-III. The rate of insulin usage is at most 37% depending on whether all insulin orders are administered. This is comparable with the rate in MIMIC-III (37%) (Fitzgerald et al., 2021) despite a slightly lower rate of hyperglycaemic episodes on day 1 of the ICU in the eICU-CRD at 29.8%, compared with 36% in MIMIC-III (Baker et al., 2020). However, eICU-CRD has a higher rate of patients with a mean blood glucose above 180 mg/dL (17.1% vs. 14%), with these differences likely related to variation in measurement frequency. For comparison, in the sicker cohort of the multicentre NICE-SUGAR study the rate of insulin use was 69.0% for the <180 mg/dL target arm (patients with expected ICU stay over 24 hours and not eating). In contrast to recommendations for ICU patients (Care, 2022; Evans et al., 2021), but comparable to MIMIC, there is high rate of SC insulin use (88.8% of insulin prescribed ICU stays). This may be partially explained by the lower illness severity of the cohort, as noted in (O’Halloran et al., 2020) nearly two-third of eICU-CRD ICU stays had no IV infusions (in line with our rate of IV dextrose usage of 32%). Relatedly, use of SC insulin injections is less resource intensive (Tran, Kibert, Telford, & Franck, 2019). Clinical recommendations and practice are constantly evolving with variation in their applicability to all patient groups. Analysis of the variation between the Surviving Sepsis Campaign guidelines and clinical practice found similar variation, with the recommendations being fully followed in 0.1% of cases (Reade et al., 2010).

In other areas related to glycaemic control, the eICU-CRD is again similar to published results. Diabetics make up 23% in MIMIC-III (Fitzgerald et al., 2021), 20% in NICE-SUGAR, comparable to the rate of approximately 22.9% in the eICU-CRD. Diabetic ketoacidosis (DKA) patients, a cohort requiring different management to standard ICU glycaemic control protocols, have previously been reported as accounting for 1.4% (IQR: 1.0-2.5%) of ICU admissions (Gershengorn et al., 2012), within range of the 2.4% rate in eICU-CRD.

However, the 1.2% hospital mortality rate of ICU admitted DKA patients is above average compared to the rate of 0.7% (range: 0.4% to 3.4%) in Gershengorn et al. (2012), possibly in line with previous evidence that eICU-CRD patient experience slightly above expected mortality rates (O’Halloran et al., 2020). In patients receiving insulin infusions the distribution of infusion rate was similar to that published for MIMIC-III, with most ICU stays receiving under 2 units of insulin per hour and a long tail of higher rates falling to near 0% at 20 units/hour (Baker et al., 2020).

Based on our analysis it would appear most eICU-CRD patients stayed in ICU for a relatively short period of time and were able to tolerate some form of oral intake (Table 1 and Figure 4D). Data quality for nutritional support was variable, with nutritional information only recorded by 36.5% of the hospitals. Generally, it wasn’t possible to determine what the oral intake specifically referred to (e.g., water or oral nutritional supplement), with it largely limited to a mL quantity and generic description (e.g., “PO”) (Appendix B). Amongst the hospitals recording nutritional information in the telehealth system the rates of EN and PN varied widely from 1-23% and 1-12% (Appendix B). Half of EN was initiated within 37 hours, with clinical guidelines recommending that EN be initiated within 48 hours for patients that cannot eat orally (Singer et al., 2019). A large percentage of patients on EN appear to be underfed (Figure 4F), assuming a 1.5-2.0 calories/mL product, and 1,500-2,000 calorie per day nutritional requirement, in line with previous research (Ridley et al., 2018). The rates of oral intake are higher (∼50%) compared to a rate of 43.6% in Fadeur, Preiser, Verbrugge, Misset, and Rousseau (2020) a group with an ICU stay greater than 3 days (Figure 4A), although as noted, what the oral intake consisted of in the eICU-CRD cohort was often ambiguous.

The associations of measures of glycaemic control and nutritional support with hospital mortality were in line with previous research. The eICU-CRD cohort displayed the characteristic U-shaped risk curve for blood glucose, with attenuation of the effect for diabetics. Glycaemic variability (as measured using the standard deviation of blood glucose (Eslami, Taherzadeh, Schultz, & Abu-Hanna, 2011)) was additionally associated with increased mortality risk. While blood glucose mean level and standard deviation are strongly associated (as shown in Figure 1H), our analysis (Figure 1J) suggests that the association between variability and poor prognosis is independent of the mean blood glucose level, as previously suggested by Meyfroidt et al. (2010). The results for nutritional support were similarly in line with previous literature (Kreymann et al., 2006). A shorter time to initiation of EN, and higher rates of feeding were both associated with better patient prognosis. We emphasise that associations are confounded by other factors such as illness severity and treatment – for instance diabetics are often treated with insulin (Figure 3F) and initiation of EN requires hemodynamic stability (Yang, Wu, Yu, & Li, 2014).

### Future directions

Hernán, Hsu, and Healy (2019) describe a taxonomy of data science tasks – descriptive analysis, predictive analysis (e.g., statistical model fitting for risk factor discovery) and causal inference. The importance of descriptive (epidemiological) research in critical care has previously been noted (Garland, Olafson, Ramsey, Yogendran, & Fransoo, 2013). It aids in understanding where variation in clinical practice and patient outcomes exist. As a large geographically dispersed population, the eICU-CRD can provide valuable information for this task. This paper initiated such research in the areas of glycaemic control and nutritional support, building upon previous descriptive analysis using eICU-CRD by O’Halloran et al. (2020) and MIMIC-III (Baker et al., 2020). Further work may expand this work to other clinical areas, or continue work on glycaemic control and nutritional support through creation of benchmark datasets for comparison of blood glucose forecasting and insulin recommendation algorithms (as done for MIMIC-III (Arévalo et al., 2021)).

As noted above there are a variety of open questions in critical care research relating to glycaemic control and nutritional support. The eICU-CRD appears to be a valuable resource for evidence generation in several of these areas, whether through replication of existing observational research or the design of novel research studies. For instance, increased glycaemic variability has been linked to poorer patient outcome (as seen in Table 1)(Ceriello & Ihnat, 2010). Most patients in the eICU-CRD have 4 or more blood glucose measures per day and detailed information on confounding factors, enabling research in this area.

Similarly, in the area of nutritional support, studies such as Zheng et al. (2022) on EN vs. no EN and patient outcome appear wholly or partially replicable in eICU-CRD. Data missingness presents one barrier to these research topics, with research focused on insulin therapy or nutritional support likely to need to exclude up to 50% of the hospitals in the eICU-CRD to have a near complete dataset.

The eICU-CRD is unique among open access critical care databases in containing many hospitals. This opens the possibility that the eICU-CRD may be quite a useful resource for observational treatment comparison research. In recent years causal inference has been given a sound theoretical basis in the work of authors such as Judea Pearl and James Robins (e.g., Pearl (2009) and Robins (1997)) leading to the generation of a wide variety of methods and software libraries for the estimation of treatment effects in complex observational datasets (e.g., targeted learning (Van der Laan & Rose, 2018)). The necessary assumption of positivity (that all in target population have some chance to be treated/untreated) may be difficult to meet in single-centre data collections where protocols (such as described in Wilson et al. (2007) for insulin) may limit the variation in who gets treated. While participation in Philips telehealth may bring some degree of standardisation, the analysis above revealed considerable variation in factors such as use of insulin and EN/PN (Appendix B). However, any such analysis would still need to adjust for confounding, which given the heterogeneity in eICU-CRD presents its own challenges.

### Strengths and limitations

This analysis is not without limitations. Ultimately certain assumptions needed to be made in extracting the relevant information from the various tables in eICU-CRD. We have released our source code documenting these assumptions. Of course, care must still be taken even where the assumptions are correct, as in all EMR collections there may be ambiguity between the data and what occurred in clinical practice. Not all clinically relevant information may have been entered into the telehealth system from which eICU-CRD is built, or it may have been entered incorrectly. Similarly, our data extraction is not exhaustive, with information such as administration of glucocorticoids not included. Further cleaning of the data may also be possible. For example, data exploration suggests it may be possible to improve the data completeness characteristics of the *medication* table orders by examining all orders for a particular patient during an ICU stay and assuming doses and frequencies repeat across orders.

## Conclusions

The eICU-CRD is a unique and valuable resource for glycaemic control and nutritional support research. Issues of data quality and completeness that we have documented will assist in assessing its suitability for addressing specific research questions. We urge that further open access EMR resources are made available to support advances in critical care and data science research.

## Data Availability

The data underlying this article are freely available at https://eicu-crd.mit.edu/ and can be accessed following completion of the required training and data usage agreements.

## Appendix A

**eicu_crd_glucose data sources and schema**

Below we describe the source tables and resulting schema of the curated *eicu_crd_glucose* schema, which can be created using the dbt project code in the eicu_glucose folder of the project Github repository.

**Figure A1.**
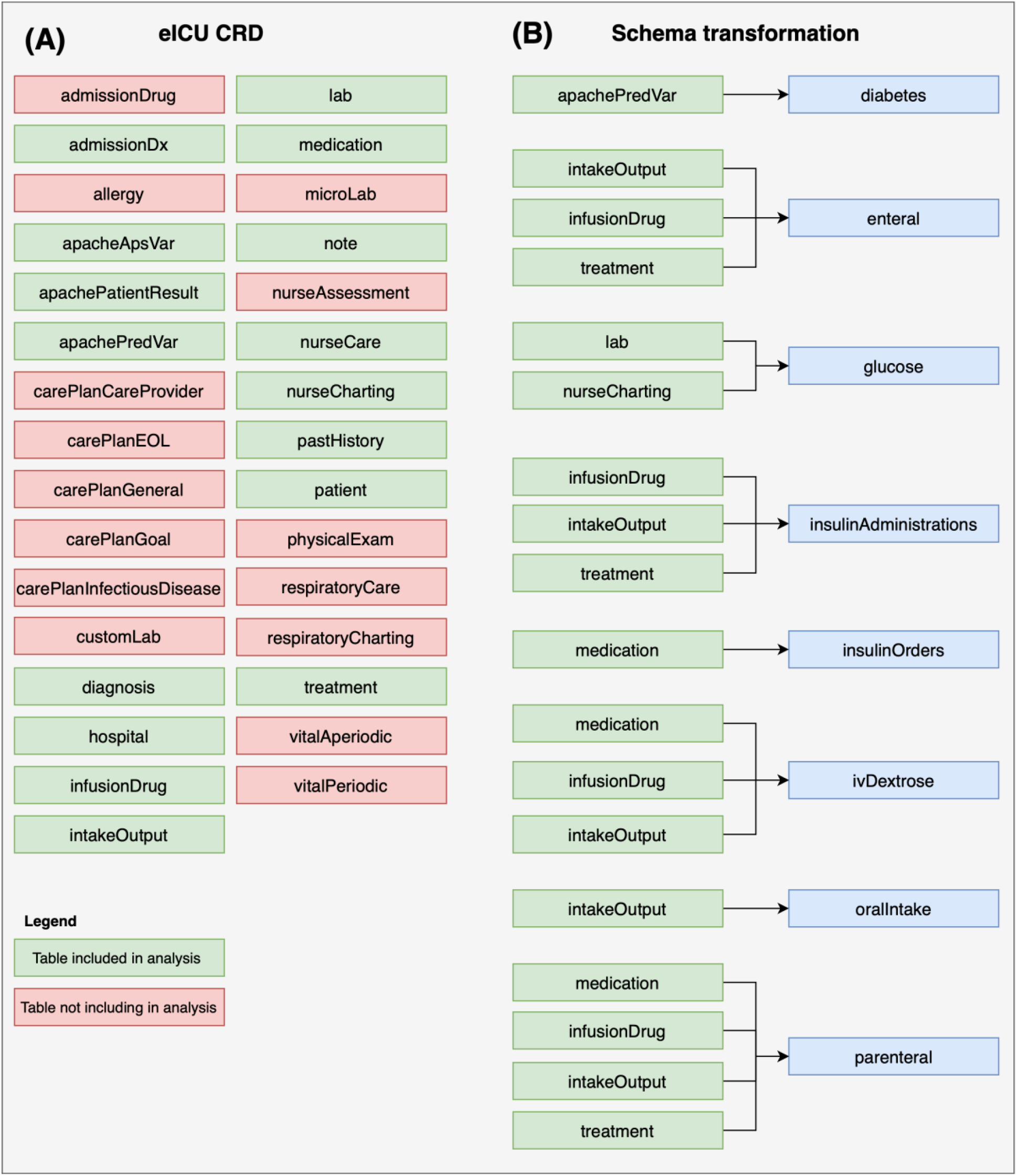
Illustration of the eICU-CRD tables included in the analysis, and the process of schema transformation to create a blood glucose management and nutrition support focused schema/database.

**Figure A2.**
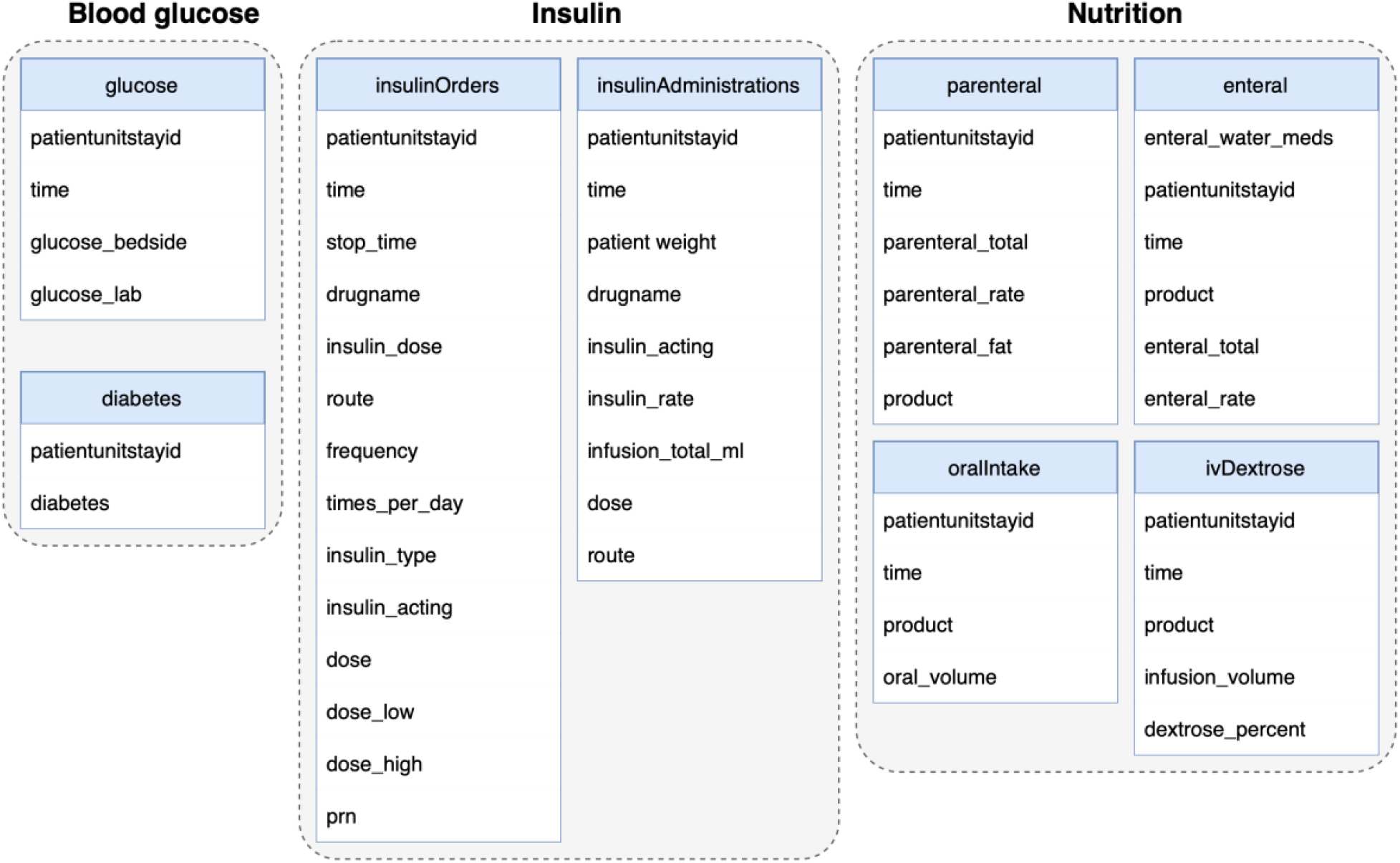
Information included in the blood glucose management and nutrition support focused schema/database.

## Appendix B

**Additional results**

Additional descriptive results

**Table B1.** Admission diagnosis and hospital outcome

|  | All | Survived hospital | Died in hospital |
| --- | --- | --- | --- |
| *APACHE Admission diagnosis (non-operative)* |  |  |  |
| *Sepsis* |  |  |  |
| Sepsis (pulmonary) | 8,862 | 7,221 (81.5%) | 1,641 (18.5%) |
| Sepsis (renal) | 5,273 | 4,682 (88.8%) | 591 (11.2%) |
| Sepsis (GI) | 2,881 | 2,302 (79.9%) | 579 (20.1%) |
| Sepsis (unknown) | 2,602 | 2,122 (81.6%) | 480 (18.4%) |
| *Cardiovascular* |  |  |  |
| Myocardial infarction | 7,228 | 6,883 (95.2%) | 345 (4.8%) |
| Heart failure | 6,617 | 5,934 (89.7%) | 683 (10.3%) |
| Rhythm disturbance | 4,827 | 4,542 (94.1%) | 285 (5.9%) |
| Cardiac arrest | 4,580 | 2,235 (48.8%) | 2,345 (51.2%) |
| Unstable angina | 2,658 | 2,613 (98.3%) | 45 (1.7%) |
| *Neurological* |  |  |  |
| Stroke | 6,647 | 5,985 (90.0%) | 662 (10.0%) |
| Coma | 2,902 | 2,642 (91.0%) | 260 (9.0%) |
| Seizure | 2,776 | 2,678 (96.5%) | 98 (3.5%) |
| ICH | 2,450 | 1,983 (80.9%) | 467 (19.1%) |
| *Metabolic* |  |  |  |
| DKA | 4,825 | 4,792 (99.3%) | 33 (0.7%) |
| *Respiratory (excl. sepsis)* |  |  |  |
| Emphysema/bronchitis | 4,494 | 4,166 (92.7%) | 328 (7.3%) |
| Pneumonia | 4,101 | 3,516 (85.7%) | 585 (14.3%) |
| Respiratory arrest | 2,599 | 2,033 (78.2%) | 566 (21.8%) |
| *Gastrointestinal/Renal (excl. sepsis)* |  |  |  |
| Upper GI bleed | 3,238 | 3,037 (93.8%) | 201 (6.2%) |
| Lower GI bleed | 2,423 | 2,307 (95.2%) | 116 (4.8%) |
| Renal failure | 2,349 | 2,151 (91.6%) | 198 (8.4%) |
| *Other* |  |  |  |
| *APACHE Admission diagnosis (operative)* |  |  |  |
| *Cardiovascular* |  |  |  |
| CABG | 4,543 | 4490 (98.8%) | 53 (1.2%) |
| Aortic valve replacement | 2,239 | 2201 (98.3%) | 38 (1.7%) |
| Carotid endarterectomy | 1,741 | 1728 (99.3%) | 13 (0.7%) |
| *Neurological* |  |  |  |
| Craniotomy | 1,193 | 1175 (98.5%) | 18 (1.5%) |
| *Gastrointestinal* |  |  |  |
| GI obstruction | 1,044 | 939 (89.9%) | 105 (10.1%) |

Characterisation of hospital variability

**Figure B1.**
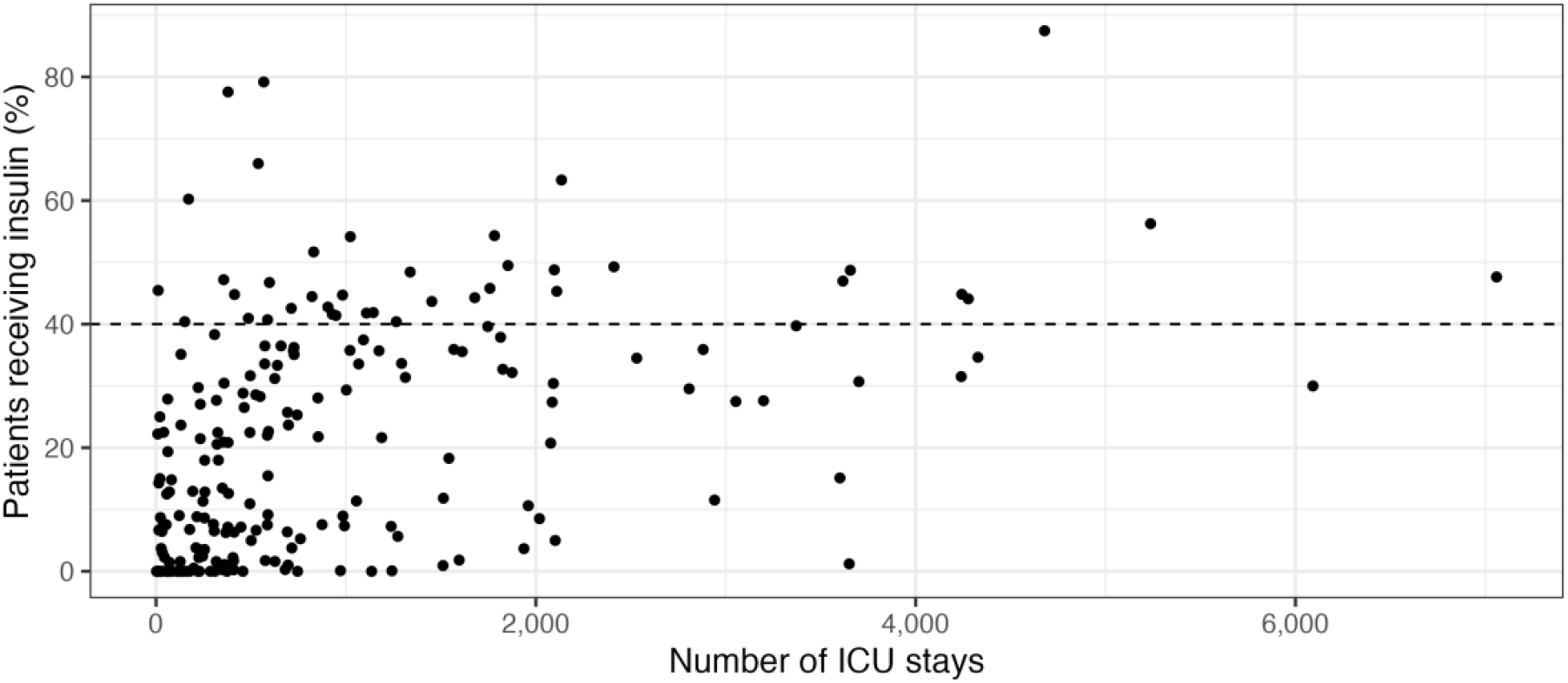
Percentage of ICU stays having an insulin order or receiving an infusion by number of ICU stays per hospital

**Figure B2.**
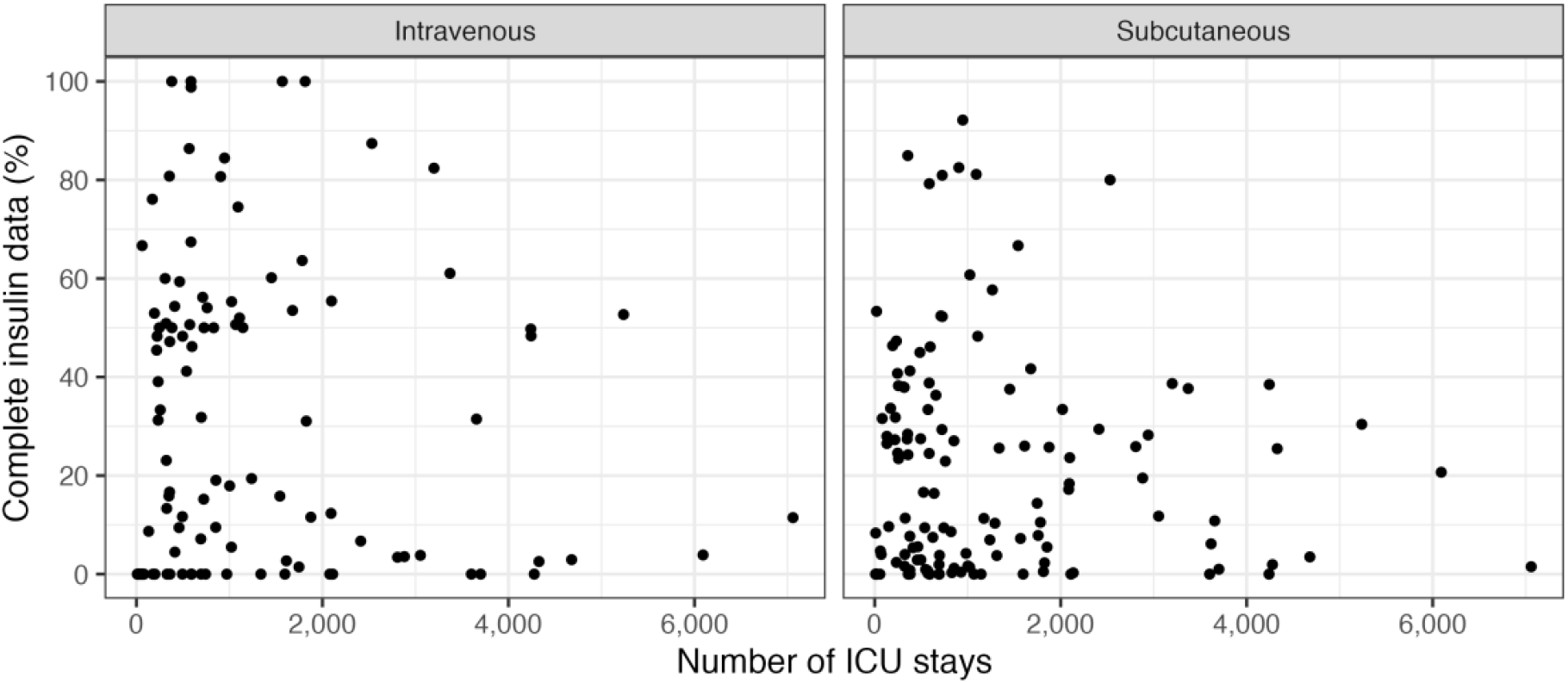
Percentage of insulin orders having complete information by number of ICU stays per hospital

**Figure B3.**
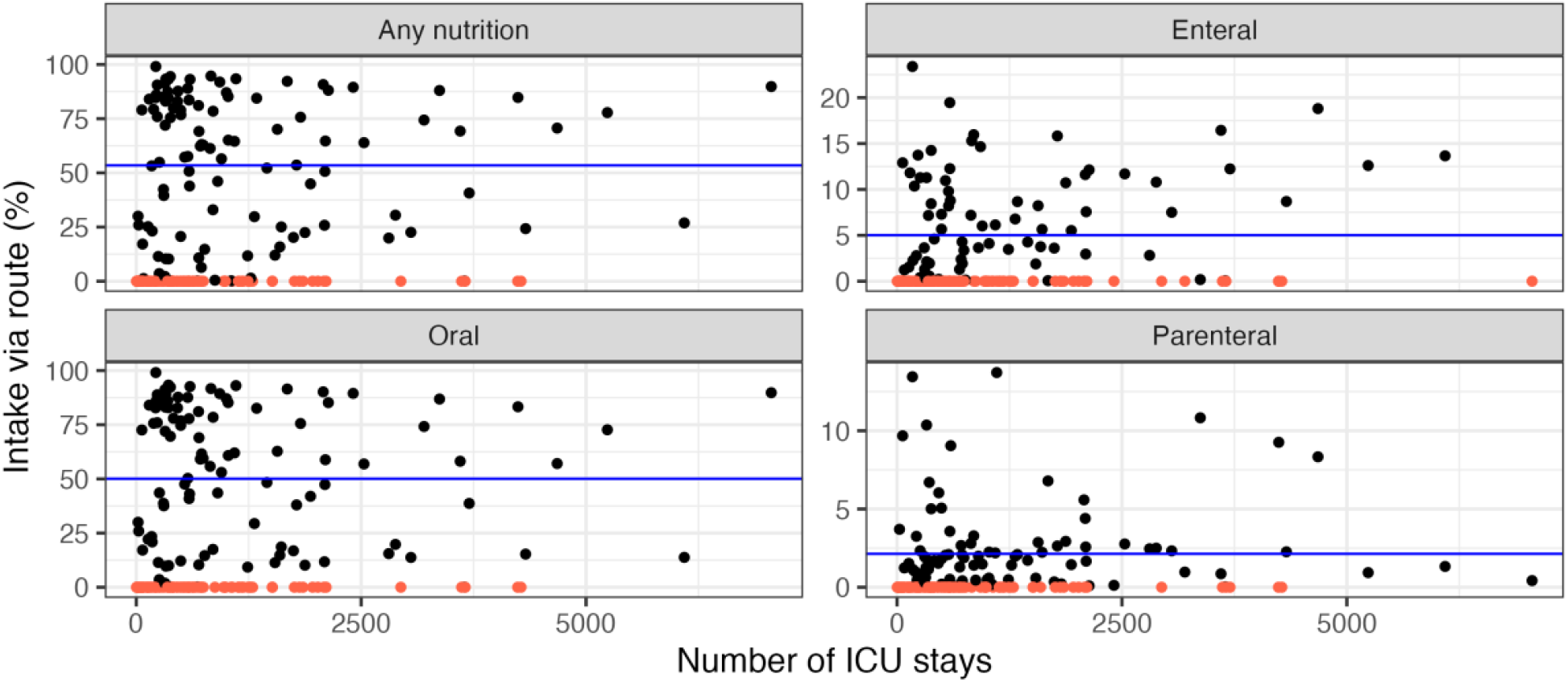
Percentage of ICU stays receiving nutritional support by number of ICU stays per hospital

## Notes

### Competing Interest Statement

The authors have declared no competing interest.

### Funding Statement

This study (via a PhD scholarship) was funded by UNSW Sydney, eHealth NSW and the Commonwealth Scientific and Industrial Research Organisation (CSIRO).

### Author Declarations

A waiver of consent that has previously been obtained from the Institutional Review Boards of MIT and BIDMC is applicable to these datasets due to their retrospective use of routinely collected EMR data. Institutional ethics for the research project was obtained from UNSW Sydney - HC220829.

